# Pediatric Diarrhea Patients Living in Urban Areas Have Higher Incidence of *Clostridioides difficile* Infection

**DOI:** 10.1101/2022.04.20.22274064

**Authors:** Ayodele. T. Adesoji, Osaro Mgbere, Charles Darkoh

**Author notes:** **Corresponding Author:** Charles Darkoh, University of Texas Health Science Center, School of Public Health, Houston, TX, USA.

## Abstract

*Clostridioides difficile* infection (CDI) is a major cause of antibiotic-associated diarrhea and an unappreciated contributor to child mortality in low- and middle-income countries where diagnosis may be difficult. There is little information about the prevalence of CDI among infants, children, and adolescents in Africa. Seventy-six samples were collected from pediatric patients presenting with diarrhea, including infants (≤ 2 years old), children (2-12 years) and adolescents (13 ≤17 years) from three hospitals between January and December 2019. Demographic data, medical history and prior antibiotic use were recorded. Toxigenic culture and PCR were used to detect and validate the presence of *C. difficile* in the samples. A total of 29 (38.7%), 39 (52.0%) and 7 (9.3%) samples were from infants, children, and adolescents, respectively. Average age of the patients was 4.4 years. Of these samples, 31 (41%) were positive for *C. difficile* by culture and were verified by PCR amplification of *C. difficile* specific genes (*tcdA* and *tcdB*). Most positive cases were children (53.3%) and infants (40.0%) with the majority of them residing in the urban areas. Forty-nine (66.2%) of the patients had no known antibiotics exposure, whereas 29.0% and 29.7% reported use of over-the-counter antibiotics at 14 and 90 days, prior to the hospital visit, respectively. CDI is relatively common among children with diarrhea in Northern Nigeria. Therefore, for effective management and treatment, more attention should be given to testing for *C. difficile* as one of the causative agents of diarrhea.

## Introduction

*Clostridioides difficile* is a strict anaerobic Gram-positive bacterium that colonizes up to 15% of healthy people [1]. It is a major cause of antibiotic-associated diarrhea worldwide, whereby antibiotic treatment reduces the abundance of competing gut microbiota, allowing *C. difficile* to proliferate and cause mild to severe diarrhea [2-4].

The toxins (enterotoxin A and cytotoxin B) released during *C. difficile* infection (CDI) cause tissue damage [5]. These two toxins, encoded by the *tcdA* and *tcdB*, are located within a 19.6 kbp pathogenicity locus. Older age, antibiotic usage, gastric acid-suppressing drugs, inflammatory bowel disease, gastrointestinal surgery, use of naso, gastric tubes, neoplastic disease, immunodeficiency, and comorbidities are risk factors for CDI [6].

The United States Center for Disease Control and Prevention (CDC) in 2019 declared *C. difficile* as one of the five ‘urgent health threats’ because of the significant risk associated with antibiotic use [7]. Despite this urgency, prevalence of CDI in low and middle-income countries is underestimated. In developing nations where self-medication occurs frequently, higher incidence of CDI may be expected. Outbreak of CDI in developed countries have increased vigilance in these countries but little is known about CDI in Nigeria [8]. Adegboyega. [9] and Doughari et al. [10] found *C. difficile* spores and the bacterium, respectively, from samples obtained from hospital environments. Oguike and Emeruwa [11] isolated *C. difficile* from 156 (48.8%) out of 320 stool samples collected from infants under the age of 5 years and confirmed occurrence of cytotoxin production from 25 (14.8%) of the isolates. Onwueme et al., [12] reported that among 71 HIV out-patients at one hospital, 10 (14.1%) and 61 (85.9%) of *C. difficile* isolates were toxin positive and toxin negative, respectively.

Normally, CDI testing in developing countries is not routine owing to the lack of resources for diagnosis and culture facilities for obligate anaerobes [13]. This is unfortunate because majority of global mortality among children in the first 5 years of their life is due to diarrhea, of which a potentially large proportion could involve CDI [14, 15]. Limited testing also translates into a low index of clinical suspicion that exacerbates the lack of effort towards diagnostic screening [16, 12] resulting in misdiagnosis and incorrect treatment. The lack of information about CDI is also problematic because most African countries are popular tourist destinations and may serve as incubators of CDI [17].

In most African countries, antimicrobials are easily accessed over-the-counter (OTC) [18-20]. There is also high incidence of community- and nosocomial-associated diarrhea among children [20, 21], but few data are available on incidence of CDI. Consequently, we initiated a survey to estimate the occurrence of CDI among pediatric diarrheic outpatients (0 - 17 years) in Katsina State, Nigeria.

## Materials and Methods

### Study population and sample collection

A total of 76 participants from three hospitals in Katsina State including Malam Mande General Hospital in Dutsin-Ma and Turai Yaradua Maternity and Children hospital and Government House Clinic in Katsina metropolis were selected for this study. The Katsina metropolis was designated as urban area, while Dutsin-Ma was designated as rural area. All the hospitals are under the management of the Katsina State Hospital Board.

Only diarrhea presenting out-patients who were reporting to these hospitals between January and December 2019, and within the age of 0 and 17 years old (infant <2 years, children 2-12 years, adolescent 13-17 years) were eligible for inclusion in the study. The case definition was based on evaluation by attending physicians, where the severity of illness was indicated by whether a child was admitted to the hospital (inpatient) or treated as an outpatient.

As a result of non-availability of information on diarrhea frequency or gravity of the symptoms available, disease severity was determined by the treatment setting of the patients [22]. Accordingly, diarrhea that required hospitalization was classified as severe, those that resolved within a few hours was considered mild, while the ones that resolved within a day was classified as moderate.

Recruitment of the patients was integrated with the services of the clinic and hospital ward under the supervision of the attending physicians who referred diarrhea patients to the microbiology laboratory. Following consent, questionnaires were administered to the patients that captured demographic data, previous antibiotics use, medical history including hospitalization, and then stool samples were collected and stored at -20°C. The samples were subsequently shipped on ice to the University of Texas Health Science Center, School of Public Health Center for Infectious Diseases, Houston, Texas, USA for analysis.

### Detection of *C. difficile* in stool samples by culture

To detect *C. difficile*, the stools were tested by toxigenic culture [23], followed by PCR of the isolates, as described previously [17]. Anaerobic condition was maintained in a Bactron 600 anaerobic chamber (Sheldon Manufacturing, Cornelius, OR, USA) using 5% CO_2_, 10% H_2_ and 85% N_2_.

### PCR analysis of bacterial pellets

DNA from the isolates was extracted using the GenElute Bacterial Genomic DNA Kit (Sigma-Aldrich, St Louis, MO, USA), according to the manufacturer’s instruction. The concentration of the extracted DNA was estimated using a NanoDrop (ThermoScientific Wilmington, DE, USA). Primer specific for toxin A and B were used to assess the presence of these genes while a 16S ribosomal marker (16S rRNA) served as a positive control for the PCR reaction [24-29, 15]. The primer sequences used were as follows: TcdA2 (F-5’AGATTCCTATATTTACATGACAATAT3’, R- 5’GTATCAGGCATAAAGTAATATACTTT3’); TNC (F-5’GAGCACAAAGGGTATTGCTCTACTGGC3’, R- 5’CCAGACACAGCTAATCTTATTTGCACCT3’); 16S rRNA (F-ACACGGTCCAAACTCCTACG, R- 5’AGGCGAGTTTCAGCCTACAA3’). OneTaq Quick-Load 2X Master Mix (New England Biolabs, IPSwich, MA, USA) was used for the PCR amplification with initial denaturation temperature of 94^0^C for 30 s, followed by 36 cycles of 94^0^C for 30s; 55^0^C for 30s; 68°C for 30s; and a final extension of 68^0^C for 5 min. Gel electrophoresis (1.0%) followed by staining with ethidium bromide and UV exposure was used to assess the results.

### Toxin detection

Cdifftox activity assay was used to detect toxin activity using 48 h culture supernatant from the isolates [30] while C. *difficile* TOX A/B II ELISA test (TechLab, Blacksburg, VA, USA) was used to detect toxin production. For the ELISA test, 200 µl of the culture supernatant was treated according to the manufacturer’s instruction.

### Statistical analysis

Data obtained from the survey were subjected to descriptive statistics using frequency runs to describe the proportional distribution of the study sample by sex, age, race/ethnicity, educational status, and area of residence. To determine the associations between the history of antibiotic use, severity of diarrhea, and presence of toxigenic *C. difficile* in the stool samples of the patients, we applied chi-square inferential test of independence. To examine the relationships between length of diarrhea and hospitalization and age of children with CDI, we carried out bivariate curve fitting models and density contour mappings to determine potential patterns (clusters) within these measures. Density ellipse probability was applied to evaluate the mass of points of the measures at *p*=0.25 and *p*=0.50. Additionally, we used Pearson’s correlation coefficient to determine the linear correlation between each of the measure and age of diarrheal patients. Based on the sample data, we built a partition-based model using the area of residence, age category and sex to determine the probability of obtaining a positive CDI test result. All statistical tests conducted were 2-tailed, and probability of ≤0.05 was used as the threshold for declaring statistical significance. Data management and statistical analyses were conducted using SAS JMP Statistical Discovery™ Software version 14.3 (SAS Institute, Cary, North Carolina, USA).

### Ethics Statement

Approval to conduct this study was obtained from the Ethical Review Committee of the Ministry of Health in Katsina State, Nigeria (MOH/ADM/SUB/1152/258). Consent from responsible guardians and ‘assent’ for those >7 years old was obtained before sample collection. Guardians of participants were informed of the purpose of the study and the fact that participation in the survey was anonymous, consensual, and voluntary.

## Results

Seventy-six stool samples were collected from patients presenting with diarrhea: 29 (38.7%), 39 (52.0%) and 7 (9.3%) infants, children, and adolescents, respectively (Table 1). There were more male (63.2%) than female (36.8%) patients.

**Table 1:**
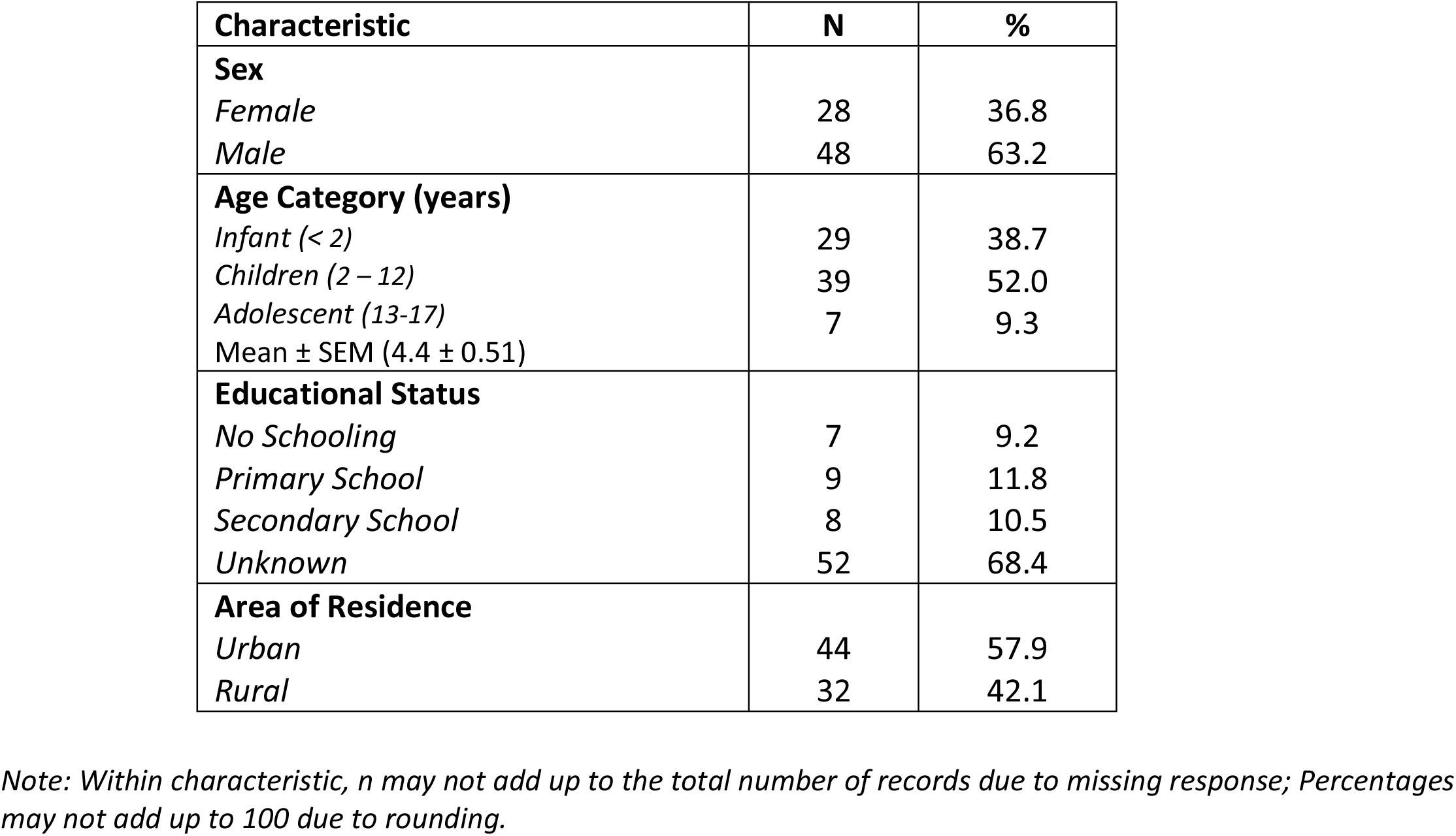
Demographic characteristics of the pediatric patients with diarrhea.

| **Characteristic** | **N** | **%** |
| --- | --- | --- |
| **Sex** |  |  |
| *Female* | 28 | 36.8 |
| *Male* | 48 | 63.2 |
| **Age Category (years)** |  |  |
| *Infant (< 2)* | 29 | 38.7 |
| *Children (2 – 12)* | 39 | 52.0 |
| *Adolescent (13-17)* | 7 | 9.3 |
| Mean ± SEM (4.4 ± 0.51) | | |
| **Educational Status** |  |  |
| *No Schooling* | 7 | 9.2 |
| *Primary School* | 9 | 11.8 |
| *Secondary School* | 8 | 10.5 |
| *Unknown* | 52 | 68.4 |
| **Area of Residence** |  |  |
| *Urban* | 44 | 57.9 |
| *Rural* | 32 | 42.1 |

*Note: Within characteristic, n may not add up to the total number of records due to missing response; Percentages may not add up to 100 due to rounding.*

For all samples, 31 (41%) grew *C. difficile* colonies while 45 (59%) were negative. All putative *C. difficile* colonies were PCR positive for *tcdA* and *tcdB*, and toxin positive by ELISA. None of the isolates that tested negative by PCR were positive for toxin production. Among the confirmed *C. difficile* patients, 11 (35%) and 20 (65%) were female and male, respectively (Fig 1a). The distributions across infants, children and adolescents were 40%, 53%, and 7%, respectively (Fig 1 b).

**Figure 1:**
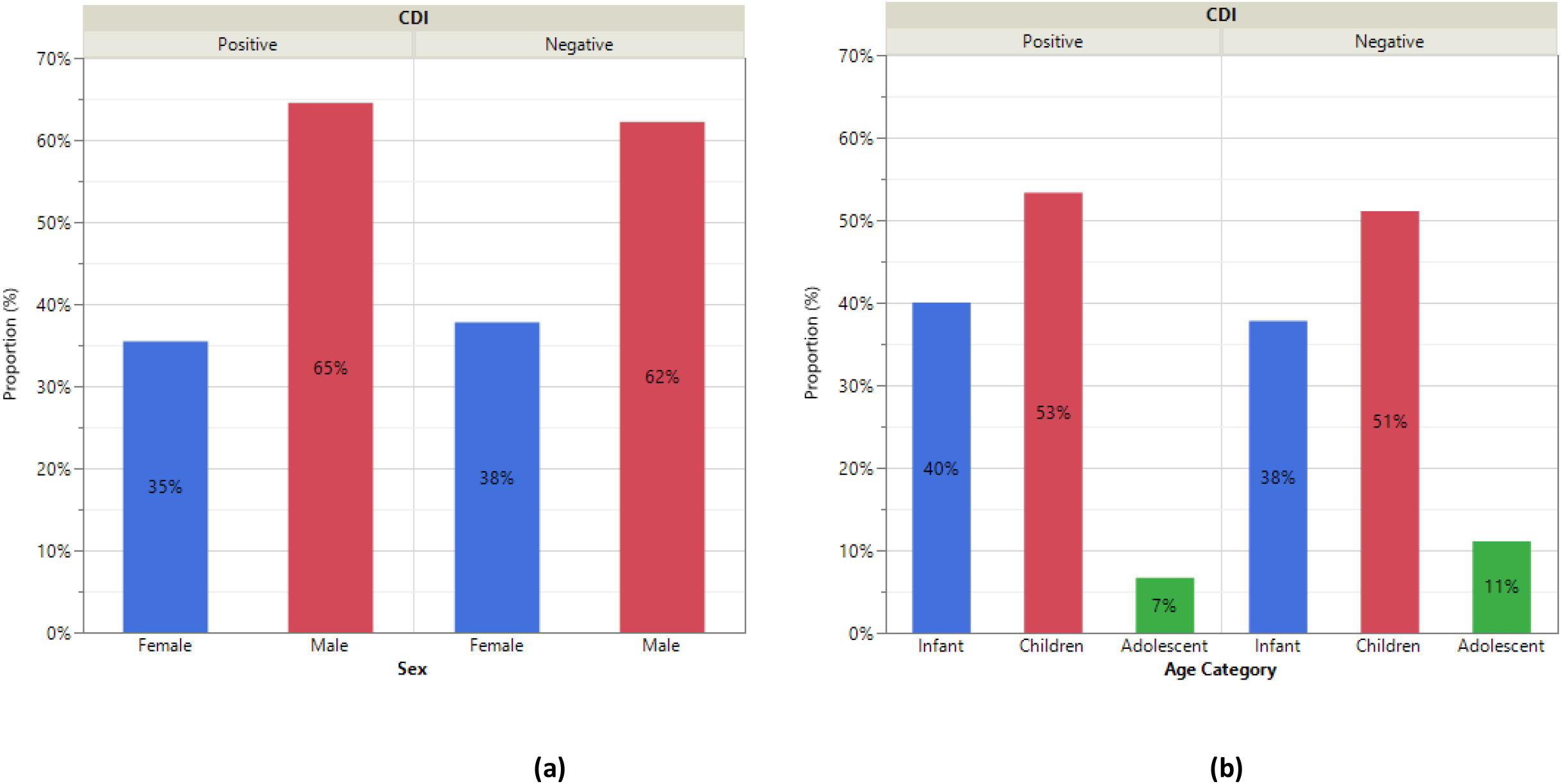
Distribution of the presence of *C. difficile* in the stools of Pediatric Patients with Diarrhea by Sex and Age Category.

The majority of the parents of the patients (66.2%) did not have any knowledge about antibiotics, whereas 29.0% and 29.7% reported OTC antibiotic use at 14 and 90 days respectively. About 13.2% and 9.2%, respectively, reported using metronidazole, and other types of antibiotics (tetracycline, ampicillin, ampiclox, cloxacilin, ciprofloxacin, gentamicin, septrin, cefuroxime, erythromycin, doxycycline, penicillin etc.). There were significant association between knowledge of antibiotics (X^2^=3.74, *p*=0.0530) and use of antibiotics in the last 14 days (X^2^=4.56, *p*=0.0328), and occurrence of CDI (Table 2). Of those who have knowledge of antibiotics (n=25, 33.8%), 18.9% of them tested positive for CDI, while 17.7% (n=11) of the patients with positive CDI indicated having taken antibiotics in the last 14 days (*p*=0.0328) prior to sample collection.

**Table 2:**
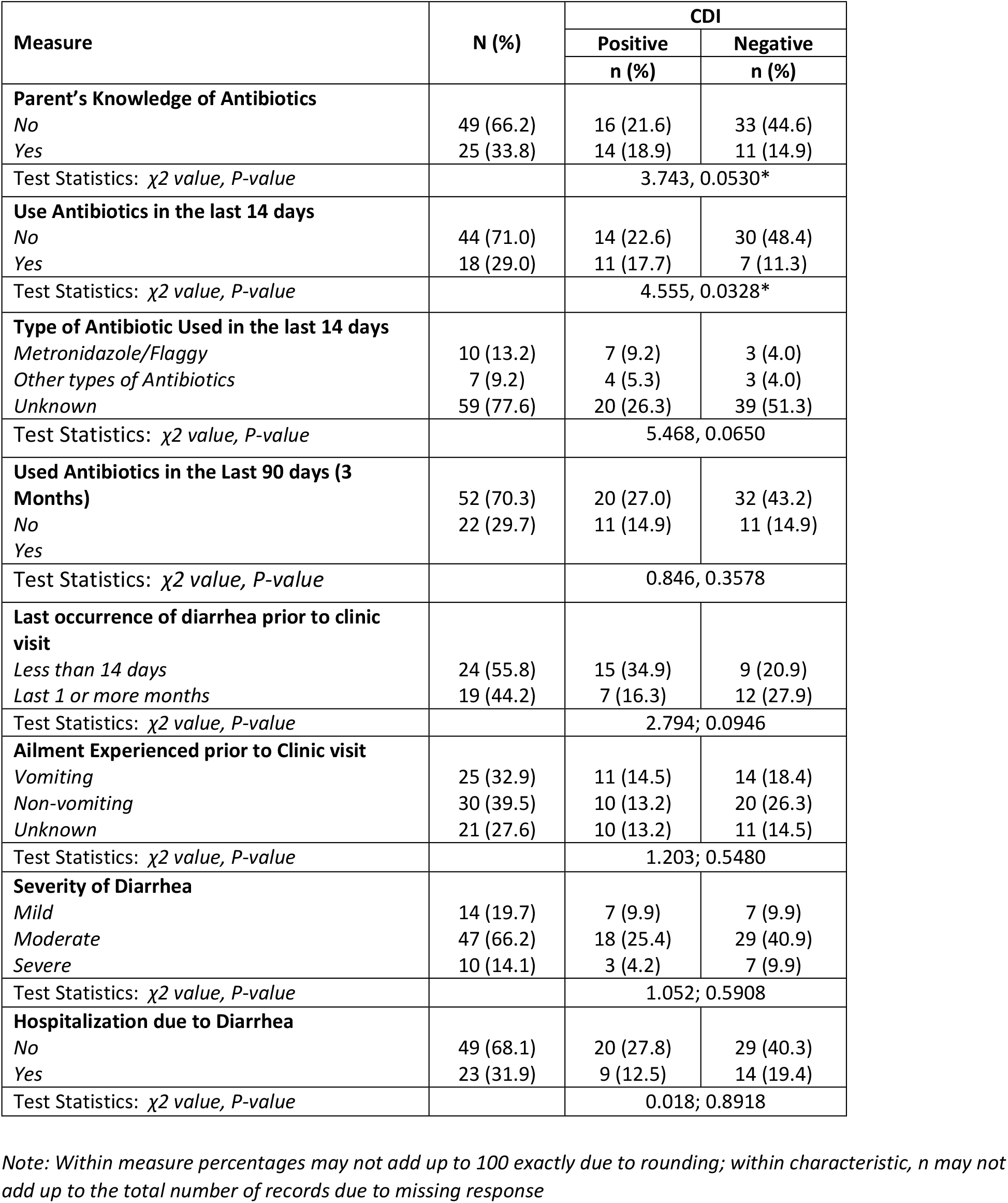
History of antibiotics used by the patients, presence of toxigenic *C. difficile*, and disease severity among pediatric patients with diarrhea.

Among the patients, 19.7%, 66.2% and 14.1% reported mild, moderate, and severe diarrhea cases, respectively. Also, 55.8 % (n=24) and 44.2% (n=19) reported suffering from diarrhea in the past 14 and 30 or more days prior to their hospital visit, respectively (Table 2). Some patients (32.9%) exhibited symptoms of vomiting prior to their visit to the clinic. Majority of the patients (68.1%) reported no prior hospitalization due to diarrhea.

Figures 2a and 2b demonstrate the relationships between length of diarrhea, hospitalization, and age of the pediatric patients. The average length of diarrhea among the patients was 5.75±1.25 days at an average age of 4.36±0.52 years, while the average length of hospitalization due to diarrhea was 7.06±3.57 days (1 week) at an average age of 3.36±0.91 years. Although the length of diarrhea and hospital stay were positively and significantly correlated (*r*=0.6870, *p*=0.0033), they were both negatively correlated with the patients’ age (*r*=-0.1946 vs. -0.1336) (Results not presented). Consequently, younger children were more likely to have longer diarrheal period and hospitalization than older ones.

**Figure 2:**
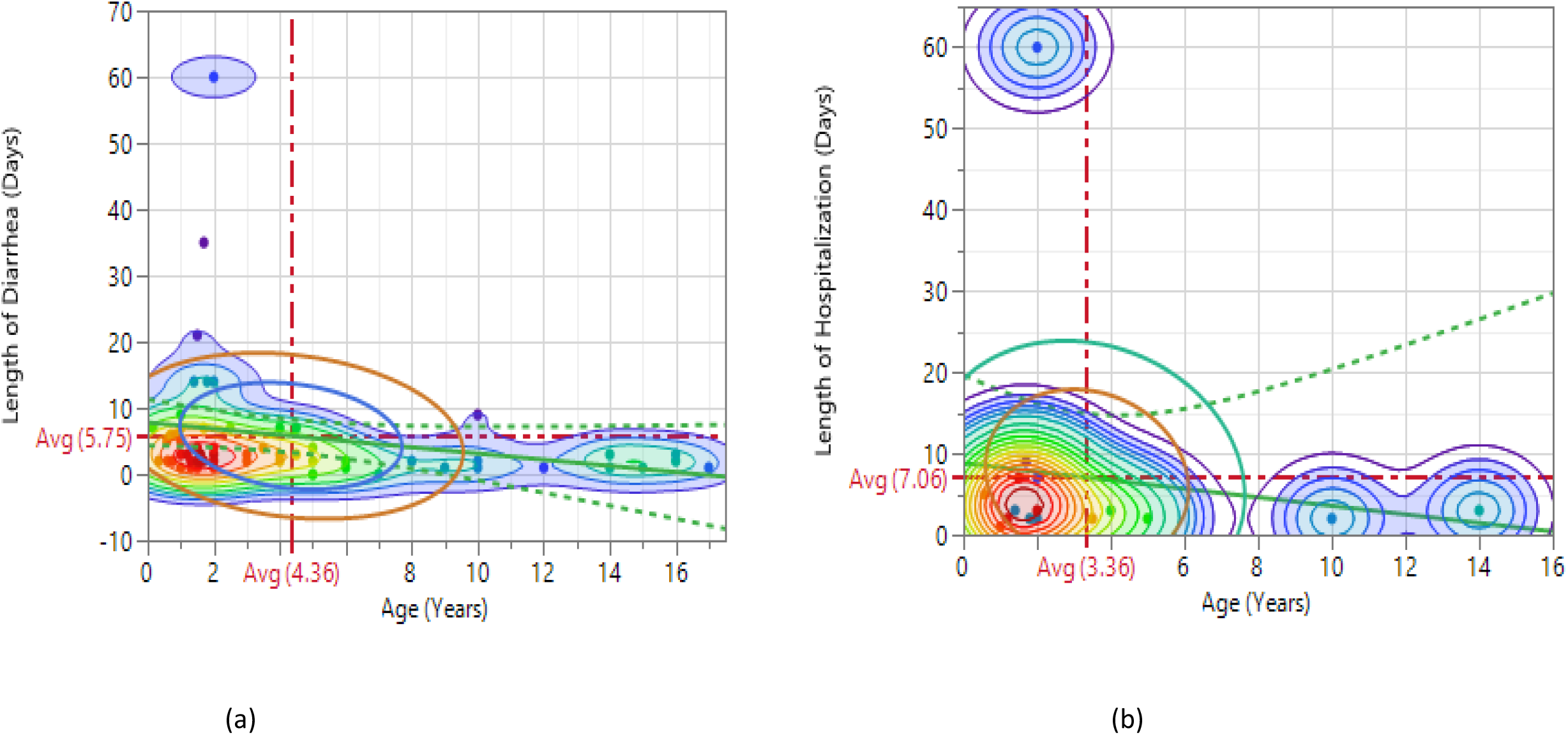
Relationship Between length of Diarrhea and Hospitalization, and Age of Pediatric Patients.

Table 3 shows the probability of obtaining a positive CDI test by selected demographic characteristics. Majority of children residing in the urban areas had more positive CDI compared to those living in the rural areas. For instance, the probability of obtaining a positive CDI among female and male children, of age 2-12 years in urban areas were 0.543 and 0.629 compared to 0.173 and 0.216 for those in the rural areas, respectively.

**Table 3:**
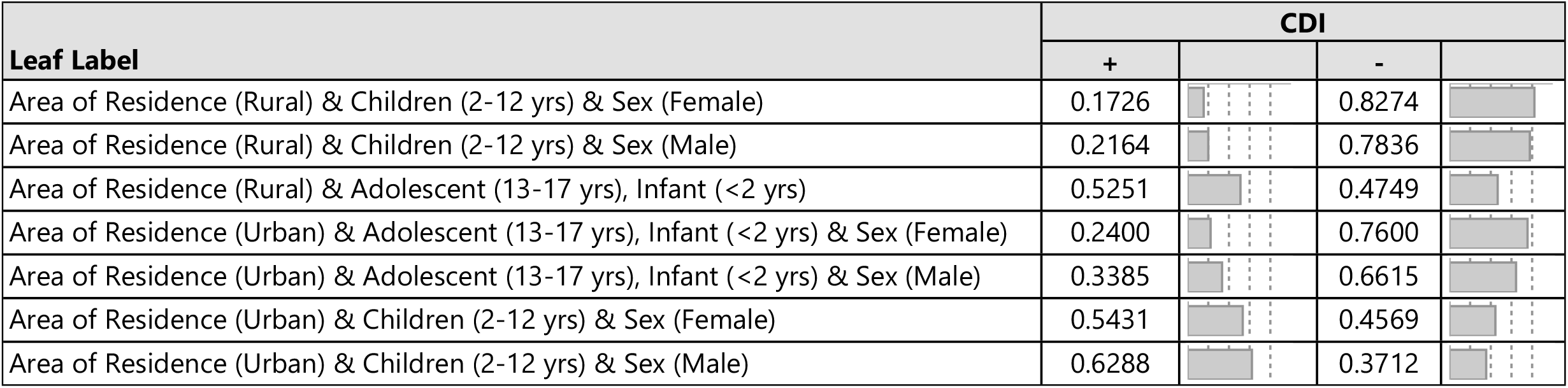
Response probability of toxigenic CDI among pediatric patients with diarrhea.

| Leaf Label | CDI |  |  |
| --- | --- | --- | --- |
|  | + |  | - |
| Area of Residence (Rural) & Children (2-12 yrs) & Sex (Female) | 0.1726 |  | 0.8274 |
| Area of Residence (Rural) & Children (2-12 yrs) & Sex (Male) | 0.2164 |  | 0.7836 |
| Area of Residence (Rural) & Adolescent (13-17 yrs), Infant (<2 yrs) | 0.5251 |  | 0.4749 |
| Area of Residence (Urban) & Adolescent (13-17 yrs), Infant (<2 yrs) & Sex (Female) | 0.2400 |  | 0.7600 |
| Area of Residence (Urban) & Adolescent (13-17 yrs), Infant (<2 yrs) & Sex (Male) | 0.3385 |  | 0.6615 |
| Area of Residence (Urban) & Children (2-12 yrs) & Sex (Female) | 0.5431 |  | 0.4569 |
| Area of Residence (Urban) & Children (2-12 yrs) & Sex (Male) | 0.6288 |  | 0.3712 |

## Discussion

CDI has been underestimated by clinicians and public health workers in Africa despite a well-documented risk of diarrhea and self-medication with antibiotics [31]. Among the 76 pediatric stool samples tested in this study, 31 (41%) were positive for *C. difficile*. This is within the prevalence rate reported by Emeruwa and Oguike [32], but slightly higher than what Plant-Paris et al. [31] reported among children less than 5 years in Kenya. Our data also showed a higher (53.33%) CDI proportion in children between 2-12 years old compared to 3-15 years old patients (30%) reported by Mutai et al., [13]. Whereas higher observation of CDI among pediatric diarrhea patients living in the urban area compared to rural dwellers of this study is contrary to what Oguike and Emeruwa [11] reported in their study. The authors reported higher (48.3 -52.5%) CDI among infants living in rural area than their urban counterparts (42-47.8%). However, in a study conducted recently in rural Ontario, Canada by Babey et al., [33], the authors noticed similarities between CDI incidence in their study compared to urban-based estimates.

We confirmed toxins A and B production in the isolates, suggesting that infants and children can be reservoirs of clinically relevant strains of *C. difficile*. The study also showed that younger children can have longer diarrheal periods and hospitalization than older children. This may be related to the immune system, since infants and children still have compromised immune systems compared to adolescents [34]. As a result, the diarrhea could be quickly resolved in adolescents than infants and children. Also, it been reported that the prevalence of *C. difficile* colonization in neonate ranges from 2% to 50% with colonization often occurring within the first week of life [35]. In contrast to adults where the rate of colonization in community and hospital settings is 3% and 20%, respectively.

Our study has some limitations that warrants cautious interpretation of the findings. First, the small sample size of participants stemmed from the fact that only diarrheal patients who reported to the hospitals were screened by the attending physicians. Consequently, this non-random sampling method placed some constraints on the generalizability of our findings, especially as some patients who had diarrhea may not have reported to the hospitals. Second, the survey was self-reported, and thus, some participants may have exhibited social desirability and recall biases in their responses. Finally, the presence of other diarrhea-causing pathogen was not tested in this study and will be the subject of our on-going research in this population.

In conclusion, attention should be given to testing for *C. difficile* as one of the causes of diarrhea during diagnosis rather than simply administrating antibiotics empirically. Testing can also promote development and implementation of strategies that regulate antimicrobials known to induce community and hospital acquired CDI. As this is the first report of CDI in northern part of Nigeria, future work will include examining the diversity of strains as well as other phenotypic and virulence-associated characteristics.

## Data Availability

N/A

## Acknowledgments

Authors would like to thank Malam Mande General Hospital, Turai Yaradua Maternity and Children hospital, and Government House Clinic for their help in recruiting patients for this study.

## Authors’ contribution

Conceived and designed the experiments: ATA, CD; Performed the experiments: ATA, CD; Contributed reagents/materials/analysis tools: ATA, CD; Data curation and statistical analysis: OM; Prepared the initial draft of the manuscript: ATA; Interpreted study findings and participate in the critical review/revision of the article for important intellectual contents: ATA, CD & OM. All authors reviewed and approved the final version of the manuscript submitted for publication.

